# RNA viromics of Southern California wastewater and detection of SARS-CoV-2 single nucleotide variants

**DOI:** 10.1101/2021.07.19.21260815

**Authors:** Jason A. Rothman, Theresa B. Loveless, Joseph Kapcia, Eric D. Adams, Joshua A. Steele, Amity G. Zimmer-Faust, Kylie Langlois, David Wanless, Madison Griffith, Lucy Mao, Jeffrey Chokry, John F. Griffith, Katrine L. Whiteson

**Affiliations:** Department of Molecular Biology and Biochemistry, University of California, Irvine, Irvine, CA, USA; Department of Biomedical Engineering, University of California, Irvine, Irvine, CA, USA; Southern California Coastal Water Research Project, Costa Mesa, CA, USA

## Abstract

Municipal wastewater provides an integrated sample of a diversity of human-associated microbes across a sewershed, including viruses. Wastewater-based epidemiology (WBE) is a promising strategy to detect pathogens and may serve as an early-warning system for disease outbreaks. Notably, WBE has garnered substantial interest during the COVID-19 pandemic to track disease burden through analyses of SARS-CoV-2 RNA. Throughout the COVID-19 outbreak, tracking SARS-CoV-2 in wastewater has been an important tool for understanding the spread of the virus. Unlike traditional sequencing of SARS-CoV-2 isolated from clinical samples, which adds testing burden to the healthcare system, in this study, metatranscriptomics was used to sequence virus directly from wastewater.

Here, we present a study in which we explored RNA viral diversity through sequencing 94 wastewater influent samples across seven treatment plants (WTPs), collected August 2020 – January 2021, representing approximately 16 million people in Southern California. Enriched viral libraries identified a wide diversity of RNA viruses that differed between WTPs and over time, with detected viruses including coronaviruses, influenza A, and noroviruses. Furthermore, single nucleotide variants (SNVs) of SARS-CoV-2 were identified in wastewater and we measured proportions of overall virus and SNVs across several months. We detected several SNVs that are markers for clinically-important SARS-CoV-2 variants, along with SNVs of unknown function, prevalence, or epidemiological consequence.

Our study shows the potential of WBE to detect viruses in wastewater and to track the diversity and spread of viral variants in urban and suburban locations, which may aid public health efforts to monitor disease outbreaks.

**Importance:** Wastewater based epidemiology (WBE) can detect pathogens across sewersheds, which represents the collective waste of human populations. As there is a wide diversity of RNA viruses in wastewater, monitoring the presence of these viruses is useful for public health, industry, and ecological studies. Specific to public health, WBE has proven valuable during the COVID-19 pandemic to track the spread of SARS-CoV-2 without adding burden to healthcare systems. In this study, we used metatranscriptomics and RT-ddPCR to assay RNA viruses across Southern California wastewater from August 2020 – January 2021, representing approximately 16 million people from Los Angeles, Orange, and San Diego counties. We found that SARS-CoV-2 quantification in wastewater correlates well with county-wide COVID-19 case data, and that we can detect SARS-CoV-2 single nucleotide variants through sequencing. Likewise, WTPs harbored different viromes, and we detected other human pathogens such as noroviruses and adenoviruses, furthering our understanding of wastewater viral ecology.

## Introduction

Municipal wastewater represents a matrix containing a wide diversity of microbes and is representative of the collective waste of a human population across a catchment area. (1). The microbial content of wastewater can be useful in determining the levels of biological contamination of an area, including the presence of human and animal feces, antimicrobial resistance genes, pathogenic bacteria, and viruses (1–6). In regard to viruses specifically, wastewater often contains high titers of bacteriophages and plant-infecting viruses, along with generally smaller proportions of viruses that infect animals – including humans (7–10). Many studies have used metagenomics to characterize the viral content of wastewater, but these studies typically rely on extracted DNA, which is unable to capture the wide diversity of RNA-based viruses (11–13). As RNA viruses can be important pathogens of humans and agricultural organisms, using metatranscriptomic sequencing to study these diverse viruses in wastewater is relevant to public health and industry and may allow for a greater understanding of the ecological processes that occur in wastewater (2, 8, 14–16).

Wastewater based epidemiology (WBE) is a useful method to detect the presence of human pathogens and may serve as an early-warning system for disease outbreaks (17). For example, WBE has been used to track the prevalence of enteric viruses such as norovirus, rotavirus, adenovirus, poliovirus, influenza, and SARS coronaviruses (18–23), with the added benefit that WBE does not rely on public health and clinical resources (17). Aside from basic detection, it has been shown that the viral load of wastewater often precedes clinical outbreaks, and may offer a forecast of the severity of localized disease outbreaks (18, 24–26). Infectious diseases are often underreported in clinical settings - likely due to asymptomatic cases, avoidance of healthcare, and incorrect diagnoses - which prevents accurate disease surveillance and incidence reporting, potentially hampering public health responses (27, 28). While not all pathogens are excreted into wastewater at detectible levels, the presence of detectible viruses in wastewater and human fecal samples indicates that WBE may still provide useful information in monitoring disease (21, 29, 30).

The coronavirus disease 2019 (COVID-19) pandemic has placed an intense strain on healthcare systems worldwide and has resulted in over 4 million human deaths (31). Caused by the 2019 emergence of the enveloped +ssRNA Severe Acute Respiratory Syndrome Coronavirus 2 (SARS-CoV-2) (32, 33), this virus has been reported in direct nasal swab patient samples, human feces, and wastewater samples (15, 23–25, 29, 34–36). While detection and quantification of SARS-CoV-2 are clearly important, the predominant method of detection, RT-qPCR, does not allow for the characterization of viral variants, which is critical to monitor during the COVID-19 pandemic to track the evolution of SARS-CoV-2 (37, 38). In light of the limitations of RT-qPCR, thousands of patient-derived SARS-CoV-2 genomes have been sequenced, and important viral variants have been discovered (37, 39–41). However, the sequencing of patient samples mainly relies on clinical samples (42), which were difficult to obtain during the COVID-19 pandemic (43, 44). As wastewater represents a composite of human waste from the total catchment area, metagenomic sequencing of these samples may allow for the detection of viral variants and pathogen diversity across larger populations and regions without further burdening healthcare workers (11, 13, 45, 46).

Given the importance of monitoring the COVID-19 pandemic and exploring RNA viral diversity, in this study metatranscriptomic sequencing and droplet digital PCR (ddPCR) were conducted in parallel to characterize the RNA viromes and viral load of SARS-CoV-2 of 94 influent samples from seven wastewater treatment plants (WTPs) representing a total population of 16 million individuals across Southern California. Through our study, we investigated several lines of inquiry: First, what is the diversity of RNA viruses across Southern California wastewater, and does viral abundance change longitudinally? Second, can we detect human-infecting viruses - including coronaviruses - in wastewater, and how does SARS-CoV-2 quantification from wastewater samples compare to county-level COVID-19 case counts? Third, can we use metatranscriptomics to detect and track the emergence of SARS-CoV-2 strain variants in wastewater over time?

## Results

### ddPCR quantification of SARS-CoV-2 and correlation with daily countywide COVID-19 cases

We quantified SARS-CoV-2 viral load in 85 influent wastewater samples from August 2020 – early January 2021 across seven WTPs in the California counties of Los Angeles, Orange, and San Diego counties. Because of COVID-19 reporting at the county level, we correlated SARS-CoV-2 N1 gene copies per liter of influent wastewater with cases within counties only (i.e. the Hyperion WTP was correlated with Los Angeles county COVID-19 cases; supplemental file SF1). Generally, the viral load of each individual WTP correlated well with the seven-day rolling average of reported countywide COVID-19 cases: Hyperion (ρ = 0.84, P < 0.001), Joint Water Pollution Control Plant (ρ = 0.83, P < 0.001), North City (ρ = 0.94, P = 0.03), South Bay (ρ = 0.37, P = 0.41), Orange County (ρ = 0.86, P < 0.001), Point Loma (ρ = 0.82, P < 0.001), and San Jose Creek (Los Angeles county, ρ = -0.15, P = 0.62) (Fig. 1).

**Figure 1:**
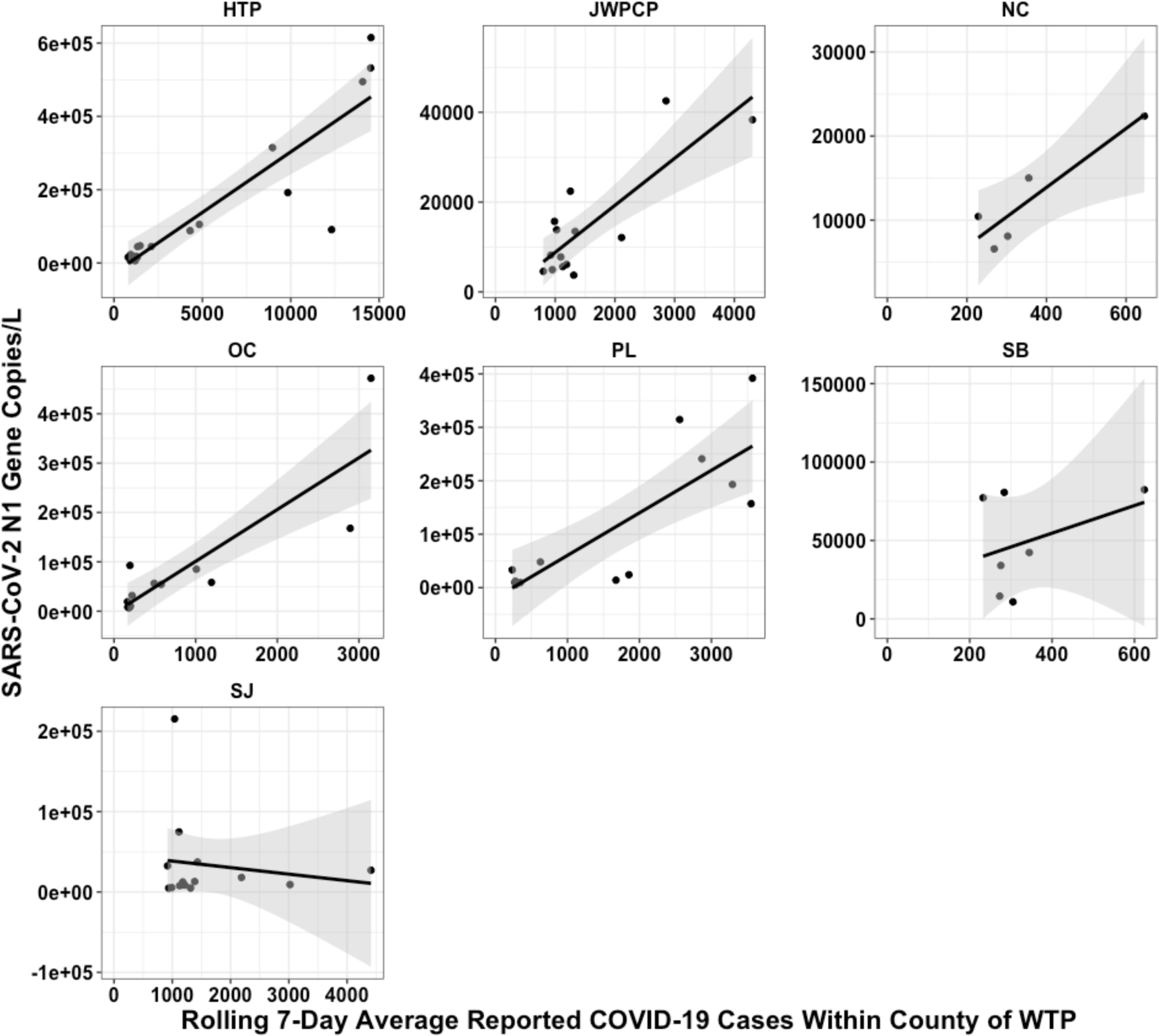
Pearson correlations of SARS-CoV-2 wastewater viral loads as measured by RT-ddPCR and the seven-day rolling average of COVID-19 case counts within county of WTP faceted by WTP. COVID-19 rolling averages and wastewater viral load were generally significantly correlated: HTP (ρ = 0.84, P < 0.001), JWPCP (ρ = 0.83, P < 0.001), NC (ρ = 0.94, P = 0.03), SB (ρ = 0.37, P = 0.41), OC (ρ = 0.86, P < 0.001), PL (ρ = 0.82, P < 0.001), and SJ (ρ = -0.15, P = 0.62).

### Sequencing library characteristics

We obtained a total of 1,119,674,084 quality-filtered deduplicated paired reads across 180 libraries (90 unique samples), and as we used two different library preparation strategies, we report the statistics separately as follows: For unenriched libraries, we obtained (794,773,512) reads (average: 84,55,037 range: 2,096,056 – 17,005,378) of which 40,512,123 (average: 430,980 [2.1%], range: 3,560 [0.05%] – 1,675,525 [13.5%]) were viruses and an average of 2.4% (range 0.06% - 25.1%) were human reads. For enriched libraries, we obtained 324,900,572 reads (average: 3,777,914; range: 1,182,078 - 14,392,582) of which 11,660,490 (average: 135,587 [4.7%], range: 2,580 [0.09%] – 484,944 [17.8%]) were viruses and an average of 2.0% (range 0.06% - 19.1%) were human reads. We were able to pair 172 libraries (86 unique samples) that had both unenriched and enriched samples successfully sequenced.

### Viral composition of wastewater and detection of selected pathogens

As enrichment changed the viral composition of wastewater samples, we analyzed the unenriched and enriched sample data separately. Unenriched samples contained sequences from 2,495 viruses, and the top ten most proportionally abundant viruses accounted for an average of 97.6% of the total abundance. The average relative abundances ± standard deviation of these “top ten” viruses were as follows: Tomato brown rugose fruit virus (66.0 ± 7.4%), Pepper mild mottle virus (10.6 ± 3.7%), Cucumber green mottle mosaic virus (10.4 ± 3.9%), Tomato mosaic virus (4.8 ± 2.9%), Tobacco mild green mosaic virus (2.1 ± 1.2%), Tropical soda apple mosaic virus (1.5 ± 0.8%), Tomato mottle mosaic virus (1.4 ± 1.8%), crAssphage (0.4 ± 0.8%), Opuntia virus 2 (0.2 ± 0.4%), and Melon necrotic spot virus (0.2 ± 0.2%) (Fig. 2).

**Figure 2:**
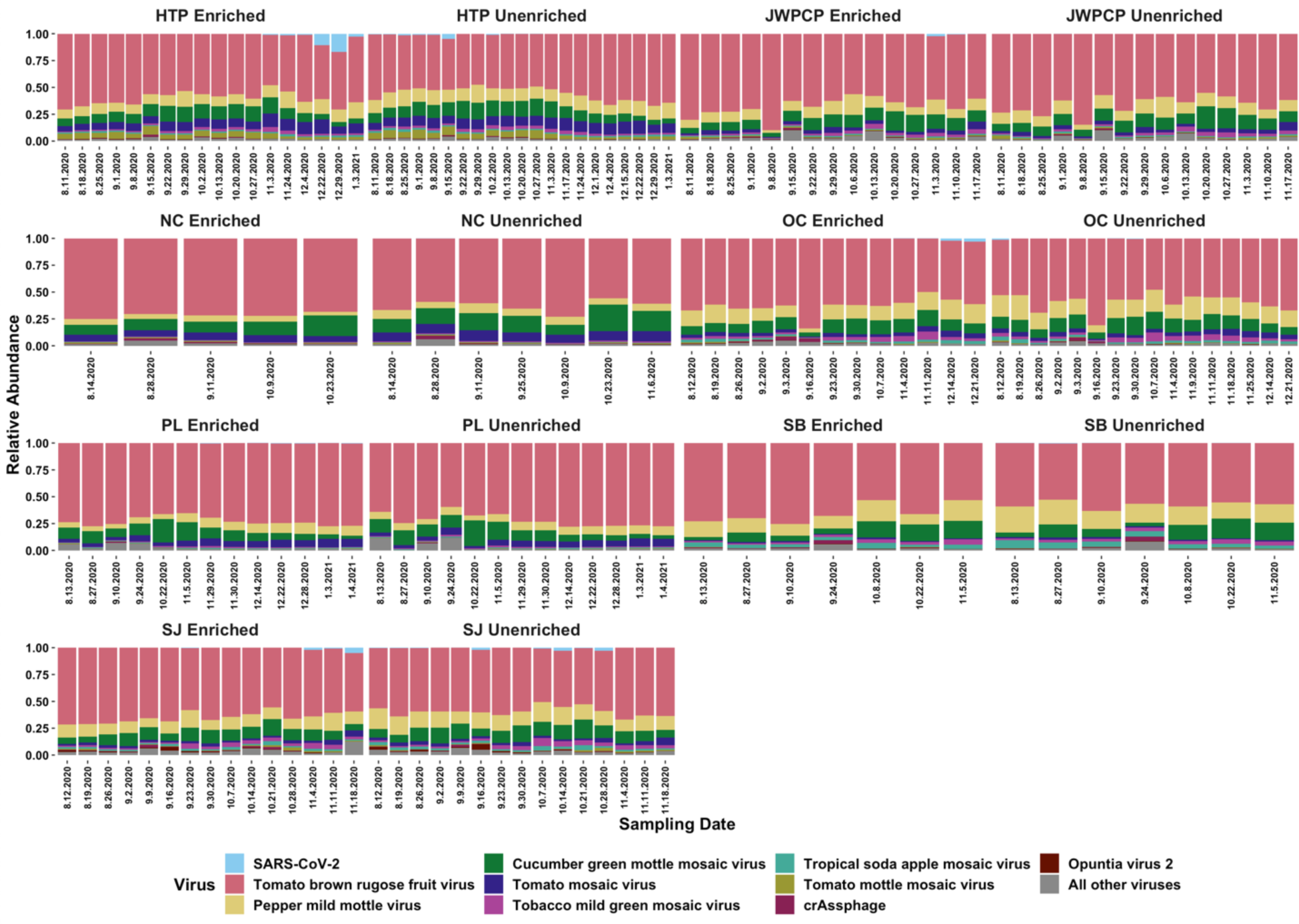
Stacked barplot indicating the ten most proportionally abundant viruses across all samples, with the relative abundance of SARS-CoV-2 being highest in HTP samples from 12/29/20. Abbreviations denote WTP, “enriched” are samples enriched for respiratory viruses, and dates across the “x” axis signify sampling date.

We analyzed the data from IRV enriched samples as above, and found sequences from 2,215 viruses, with the top ten most proportionally abundant viruses accounting for an average of 97.2% of total abundance. The average relative abundances ± standard deviation of these “top ten” viruses were as follows: Tomato brown rugose fruit virus (60.2 ± 8.6%), Pepper mild mottle virus (13.0 ± 4.4%), Cucumber green mottle mosaic virus (11.5 ± 4.3%), Tomato mosaic virus (5.0 ± 3.0%), Tobacco mild green mosaic virus (2.6 ± 1.6%), Tropical soda apple mosaic virus (1.8 ± 1.4%), Tomato mottle mosaic virus (1.5 ± 1.4%), SARS-CoV-2 virus (0.9 ± 2.3%), crAssphage (0.4 ± 0.7%), and Opuntia virus 2 (0.3 ± 0.6%) (Fig. 2). We were able to detect many more reads of SARS-CoV-2 with respiratory virus enrichment, as there were only 337 SARS-CoV-2 reads (an average proportional abundance of 0.0004%) in unenriched samples, while across enriched samples, we detected 124,135 SARS-CoV-2 reads.

The Illumina Respiratory Virus Oligo Panel enriches for 40 viruses (including SARS-CoV-2), so we were able to compare viral detection in 86 enriched and unenriched samples, along with two non-respiratory viruses often detected in wastewater: norovirus and PMMoV. In enriched libraries, we detected the presence of SARS-CoV-2 in 68 samples, Human coronavirus (HCoV)-OC43 in 22 samples, HCoV-229E in one sample, Influenza A in 10 samples, human adenoviruses in 15 samples, human bocaviruses in 13 samples, and noroviruses in 52 samples. In unenriched samples, we detected SARS-CoV-2 in 24 samples, HCoV-OC43 in two samples, and noroviruses in 59 samples. Likewise, we detected PMMoV in all samples regardless of enrichment (Fig. 3), indicating that sequencing was successful.

**Figure 3:**
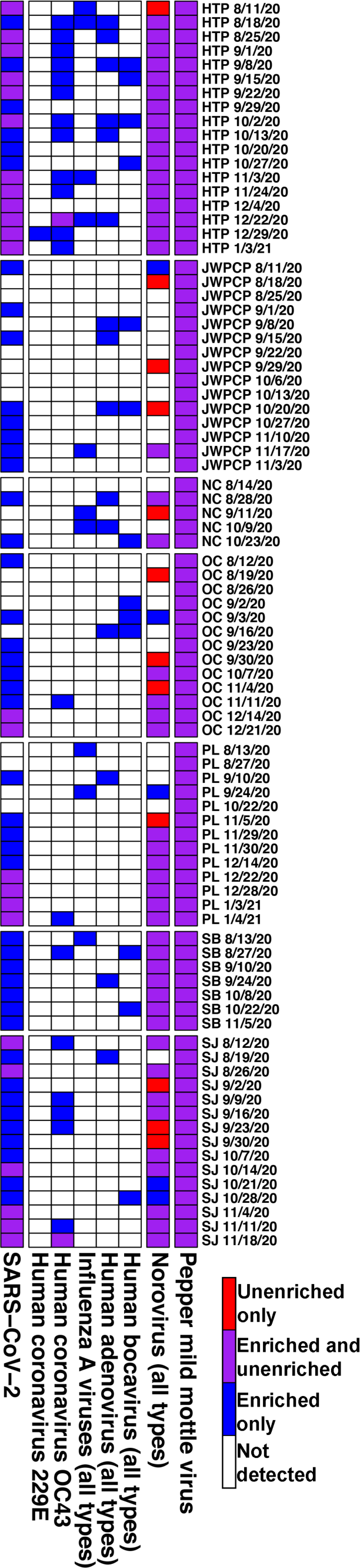
Heatmap indicating the presence or absence of reads mapping to each listed virus. Heat color denotes if a virus was detected in unenriched samples, respiratory virus enriched samples, both, or neither type of sample, and abbreviations signify WTP.

### Wastewater viral ecology

We analyzed the Shannon indexes for unenriched samples and found that overall alpha diversity was significantly different between wastewater sampling sites (F(_6,87_) = 7.5, P < 0.001), then used Tukey’s HSD post-hoc pairwise comparison testing to show that only NC-HTP, PL-HTP, PL-JWPCP, PL-OC, and PL-SJ alpha diversities were different from each other (P_adj_ < 0.05, Fig. 4). We also compared the Bray-Curtis dissimilarities of the samples with Adonis and found that overall treatment plants’ beta diversity was significantly different (R^2^ = 0.41, P < 0.001, Fig. 4). Additionally, we used ANCOM to test for differential abundance of viruses at > 0.0001 average relative abundance between treatment plants. This resulted in 16 viruses being differentially abundant between treatment plants (W > 46, Padj < 0.05 each, Fig. 4). In enriched samples, we compared the proportional abundances of human respiratory viruses at > 0.0001 average relative abundance and show that SARS-CoV-2 and HCoV-OC43 were significantly different across treatment plants (W > 46, Padj < 0.05 each, Fig. 4).

**Figure 4:**
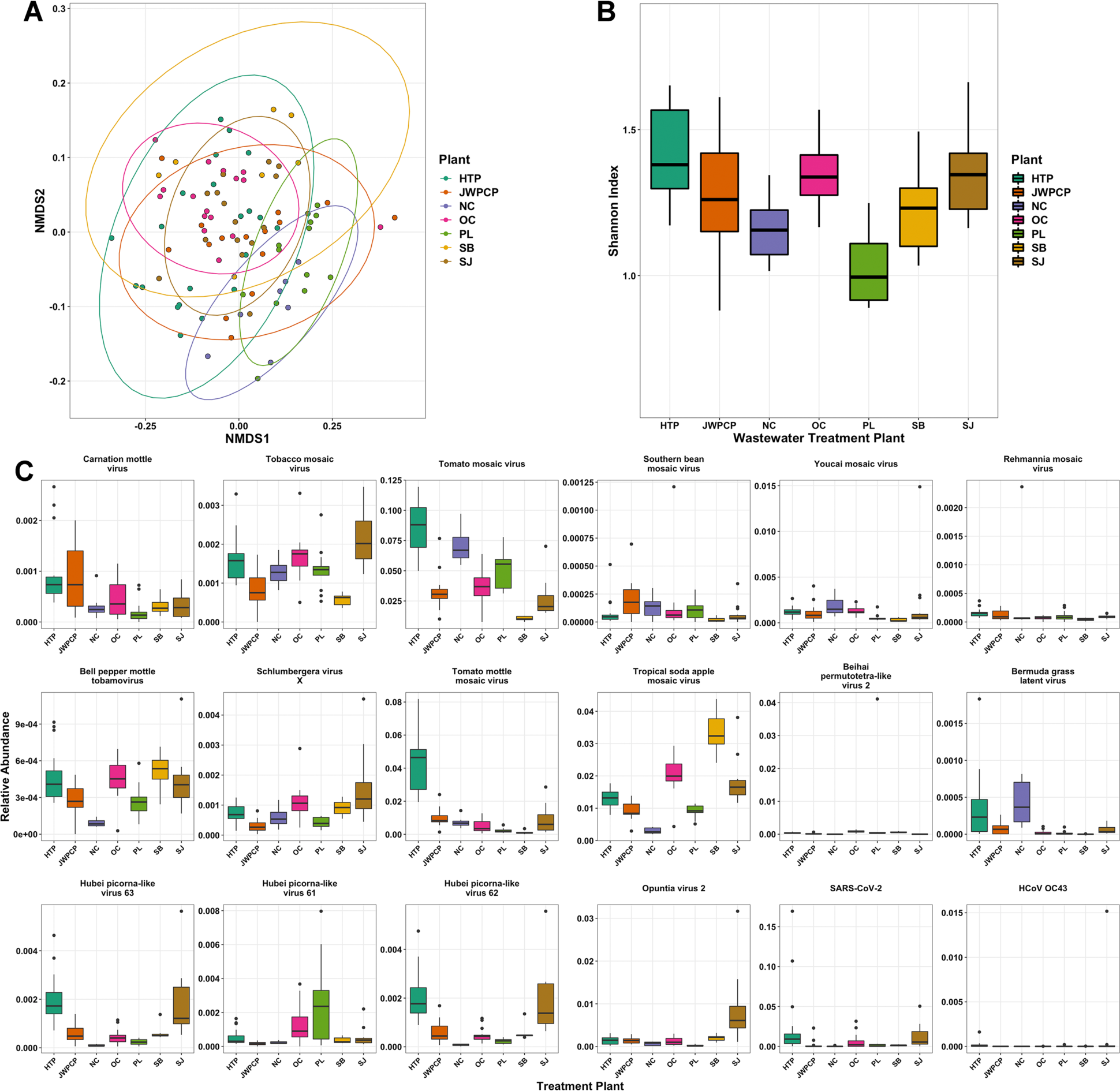
A) Nonmetric multidimensional scaling (NMDS) ordination of the Bray-Curtis dissimilarities of all unenriched samples colored by WTP. Treatment plants were significantly different from each other (R^2^ = 0.41, P < 0.001). B) Boxplots of the Shannon diversity indexes colored by WTP. Overall alpha diversity was significantly different (F_(6,87)_ = 7.5, P < 0.001). C) Boxplots of virus relative abundances that significantly differed between treatment plants as tested with ANCOM, with the right bottom two panels (SARS-CoV-2 and HCoV OC43) representing enriched samples *only*.

As we sampled the treatment plants multiple times over our study (up to 145 days from first sample), we were able to study how the viromes changed longitudinally. We compared diversity measures over time with LMEs using treatment plant as a random effect and show that both alpha and beta diversity remained stable across the sampling periods in unenriched samples (Shannon: t = 0.21, P = 0.84, Bray-Curtis: t = -0.31, P = 0.76). We also used LME on viruses present at > 0.0001 average relative abundance with treatment plant as a random effect and showed that 19 viruses’ relative abundances changed over time (P_adj_ = < 0.05, Fig. S1). We specifically were interested in SARS-CoV-2 in enriched samples and used LMEs with treatment plant as a random effect, and found that the relative abundance of this virus increased over August 2020 – January 2021 (t = 4.0, P < 0.001, Fig. S1).

### Sequencing SARS-CoV-2 SNVs in wastewater samples

We were interested in reads mapping specifically to the SARS-CoV-2 genome as a way to detect single nucleotide variants (SNVs) in wastewater. Across the 68 enriched samples that had detectible SARS-CoV-2 reads, we obtained an average breadth of genomic coverage of 24.0% (range 0.2% - 99.8%) per sample at an average sequencing depth of 8.1 reads per base (range 0.002 – 177.6) (Fig. 5 and supplemental file SF2). After masking the likely problematic nucleotide sites (as suggested by https://virological.org/t/masking-strategies-for-sars-cov-2-alignments/480/14, March 2021 update), we obtained 2558 SNVs (2002 unique) across all samples, however due to the low breadth of coverage in many samples, many of these sites may be spurious or unresolved. After applying a more stringent cutoff of 50% breadth of coverage across the SARS-CoV-2 genome, we obtained 2060 SNVs (1656 unique) across 14 samples (Fig. 5 and supplemental file SF3).

**Figure 5:**
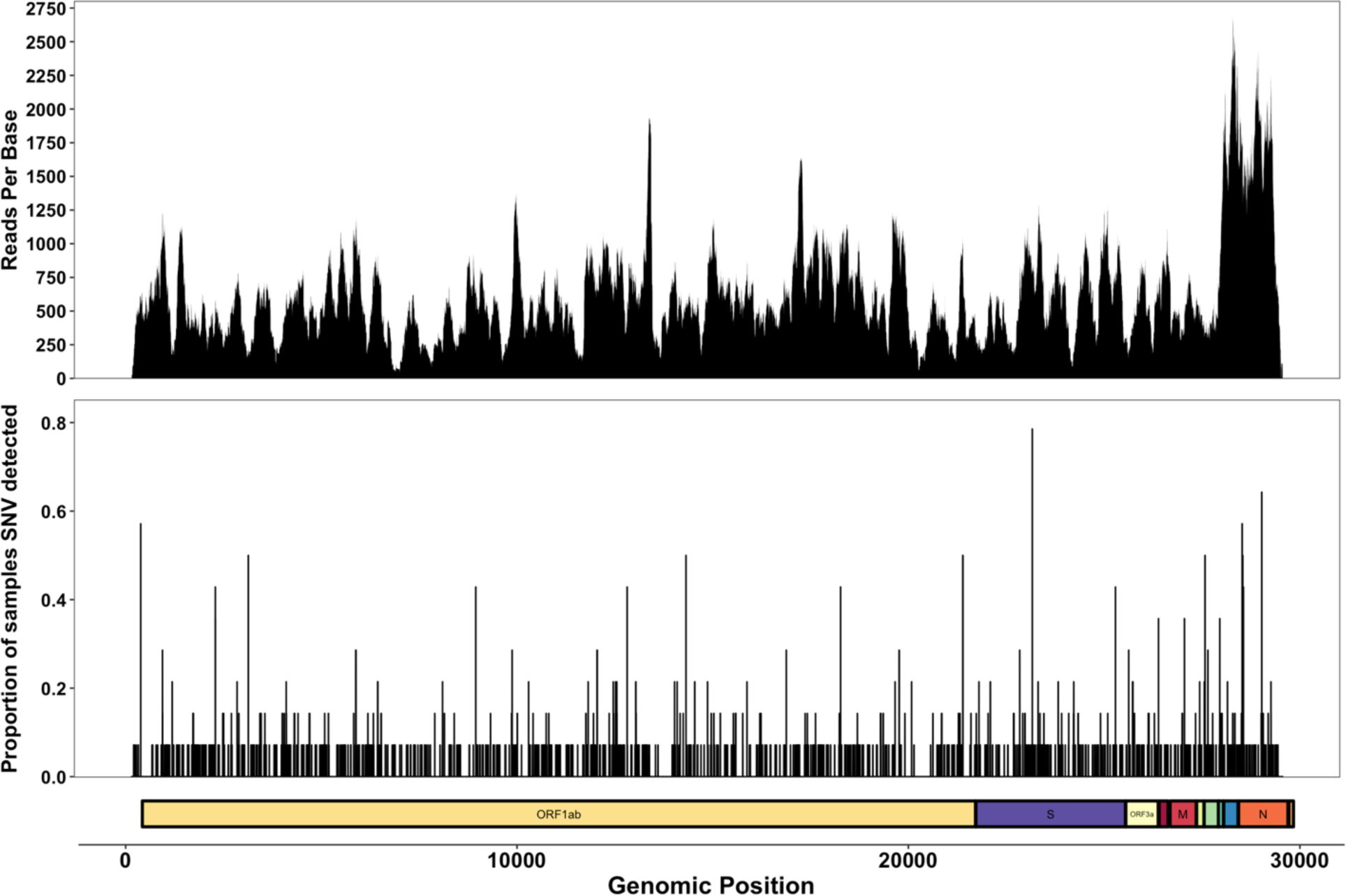
Multipanel figure showing sequence reads per base across all enriched samples, the position and proportion of SNVs across samples with >50% genomic breadth only, and an ORF map of the SARS-CoV-2 genome.

As we took samples at multiple time points per treatment plant, we were able to track the proportion of SNVs per nucleotide position over time, most notably through samples taken from the Hyperion (HTP) facility, likely due to higher viral load leading to higher sequencing depth. Within HTP, we plotted the proportion of SNVs (as compared to the reference strain) over time at nucleotide positions obtained from at least three samples with greater than 50% breadth of genomic coverage. Three of the detected SNVs are apparently fixed in the viral population (sites 241, 14408, and 23403 are all 100% SNV), while the other 17 SNVs appear to vary widely over the sampling dates (overall average %CV = 103%, range 0 – 161%) with no apparent directionality (Fig. 6).

**Figure 6:**
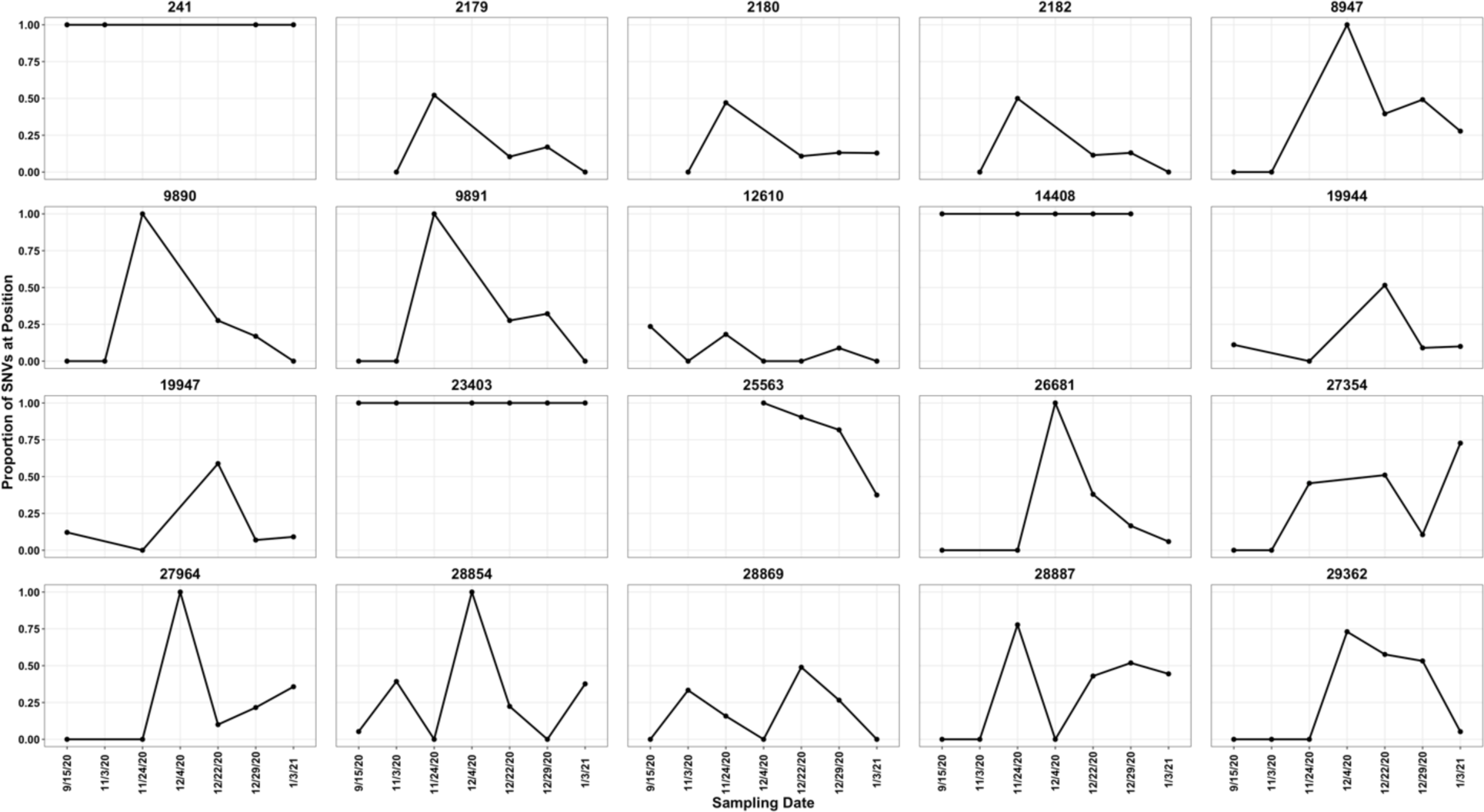
Line and dot plot of SNVs sequenced at least three times at the Hyperion WTP faceted by SARS-CoV-2 genomic position over time in samples with > 50% sequencing breadth. “Proportion of SNVs” refers to the proportion of SNVs versus the SARS-CoV-2 reference strain (i.e. 1 SNV and 3 reference reads = 0.25 proportion).

## Discussion

Our composite wastewater samples contained a diversity of RNA viruses - mainly plant-infecting viruses along with lower relative abundances of animal-infecting viruses - that differed between wastewater treatment plants (WTPs), supporting previous studies indicating that location affects the presence of viruses (13). Furthermore, several individual viruses varied over time while overall diversity remained unchanged, likely due to localized infections within WTP catchment areas (3, 9, 47). We detected several human pathogenic viruses across all WTPs, supporting the hypothesis that wastewater-based epidemiology (WBE) has the potential to inform public health and researchers about viral presence and distribution without relying on standard healthcare practices (17). Through respiratory-virus-enriched library preparation and sequencing, we detected the presence of SARS-CoV-2 at every WTP, along with several SNVs across the SARS-CoV-2 genome. This suggests that WBE can reveal the pool of potential viral variants across large geographic areas, again without adding stress to healthcare systems, with the benefit that composite sampling can collect wastewater 24 hours per day (45, 46, 48). Lastly, we show that SARS-CoV-2 viral load at WTPs generally correlates with county-level COVID-19 case counts, indicating that WBE can be useful in monitoring the severity and dynamics of disease spread (18, 19, 23–25, 49).

The vast majority of viruses present in our samples were plant viruses, mainly those infecting tomatoes, peppers, and cucumbers in the genus *Tobamovirus*. This result is consistent with other studies, which suggests that these viruses are diverse and widespread in wastewater, likely originating from agricultural runoff or human feces (3, 7, 50). Even though these viruses were ubiquitous throughout our samples, WTPs had significantly different overall viromes, indicating there may be signatures of location and wastewater catchment throughout Southern California, which has been suggested from other sampling locations (13). Aside from overall diversity, several plant- or arthropod-infecting viruses were differentially abundant between WTPs, possibly due to differences in peoples’ diet (and thus viral excretion), infected plant growth, and localized infections of arthropods (i.e. Hubei picorna-like viruses and Beihai permutotetra-like viruses) (3, 9, 47). Likewise, we found that several viruses’ relative abundances varied over time, regardless of treatment plant, suggesting that there are infection/subsidence dynamics or seasonal trends as previously suggested (7, 10, 49, 51). As the day-to-day relative abundance of viruses varied, we suggest that future wastewater surveys incorporate longitudinal and composite sampling to accurately capture viral diversity.

We detected several viruses pathogenic to humans across WTPs, including noroviruses adenoviruses, bocaviruses, coronaviruses, and influenza A - which agrees with other studies - indicating that WBE is robust and applicable to multiple viruses (18–23). By applying metagenomic or metatranscriptomic sequencing to wastewater, we can simultaneously detect a diversity of viruses, potentially alerting public health to unknown or underreported infections or new viral strains (27, 28, 46, 52). As we prepared sequencing libraries using two methods (unenriched and Illumina Respiratory Virus Oligo Panel enriched shotgun metatranscriptomics), we could compare the effects of enrichment on virus detection, and report that viral enrichment greatly improved our ability to detect influenza A and coronaviruses (especially SARS-CoV-2). We suggest that if researchers and the public health community are specifically interested in respiratory viruses, enrichment and the proper wastewater concentration /extraction methods should be employed for appropriate sensitivity and specificity to viruses of interest (23, 25, 34, 46, 53). Alongside sequencing, we also used ddPCR to quantify SARS-CoV-2 in wastewater and show that our results generally correlated well with county-reported 7-day rolling average COVID-19 cases which agrees with previous studies (23, 24, 54). We note that SARS-CoV-2 viral load and case counts were not significantly correlated at every WTP indicating that there is likely unknown variability in the survival of RNA within wastewater, or that there may be variable influent flow or water quality affecting the viral load at specific WTPs (55, 56). We also note that our comparison is limited to county-level data, rather than being a comparison of the areas contributing to the influent stream and that this could be obscuring the relationship to case data at some treatment plants.

Our respiratory-virus-enriched metatranscriptomic sequencing detected the presence of SARS-CoV-2 at every WTP - and in most of our samples - although we note that could not accurately quantify viruses through sequencing. Instead, the power of metatranscriptomic sequencing lies in our ability to detect SNVs across the SARS-CoV-2 genome (46), and depending on the sample, we often sequenced >50% of the genome at appreciable read depth. For example, we detected the GISAID clade GH (lineage B.1*) markers 241C>T, 3037C>T, 14408C>T, 23403A>G, and 25563G>T at 100% prevalence where those regions of the genome were sequenced (39, 57, 58). Likewise, as we detected over 1000 SARS-CoV-2 SNVs across our samples, we found many SNVs of putatively unknown function that have been detected in patient samples, such as 6285C>T and 9891C>T (found in, but does not solely define, variants B.1.525 and B.1.1.318 respectively), 28854C>T and 28887C>T (40, 59, 60). We also detected many SNVs also found in wastewater samples from Northern California (46) and SNVs that to the best of our knowledge, have yet to be sequenced (41, 46, 48). As our sequencing data are not quantitative, our study suggests that sequencing wastewater is useful for SNV detection across wide catchment areas, but not the true prevalence of SNVs (46). Furthermore, we recognize that wastewater likely represents a collection of different viruses’ RNA rather than one intact virion, as SARS-CoV-2 may be relatively fragile in wastewater (55, 61). Lastly, we note that most SNVs came from samples with the highest number of SARS-CoV-2 reads, and suggest sequencing samples deeply to obtain a useful breadth and depth of viral genomes for variant detection (46, 62).

## Conclusion

Wastewater-based epidemiology has the potential to aid public health in monitoring the spread and severity of disease outbreaks. Our research contributes to the growing field of WBE by showing that Southern California wastewater harbors a diversity of RNA viruses, and that these viral populations vary over time and between WTPs. Likewise, we are able to detect human viral pathogens without increasing the burden on local healthcare systems, further supporting the benefit of WBE. Through ddPCR and metatranscriptomic sequencing, we were able to measure the viral load of SARS-CoV-2 in wastewater and identify potentially novel SNVs which may assist in monitoring viral evolution and the emergence of new variants. We suggest that future researchers employ longitudinal metatranscriptomic sequencing on wastewater samples to further understand the spread of RNA viruses and how these viruses change over time.

## Materials and Methods

### Sample collection

We collected 94 1-liter 24-hour composite influent wastewater samples at seven WTPs across Southern California between August 2020 – January 2021 (Supplemental file SF4). The samples were aliquoted into 50mL tubes, then stored at 4°C until sample processing. Note that extractions were performed independently for ddPCR quantification and viromic sequencing.

### Wastewater sample processing for SARS-CoV-2 ddPCR quantification

We prepared influent samples for ddPCR following the method described in Steele et al., 2021 (63). Briefly, we first added bovine coronavirus vaccine (BoCoV, Bovilis Merck & Co, Kenliworth, NJ) to 20 mL of wastewater as a sample processing control to assess viral RNA recovery. We then added MgCl_2_ to a final concentration of 25mM and adjusted the pH to < 3.5 with 20% HCl on a mixed cellulose ester membrane (Type HA: Millipore, Bedford, MA) in replicates of six. We then transferred the HA filters to pre-loaded 2 mL ZR BashingBead Lysis tubes (Zymo, Irvine, CA), and bead beat the samples with a BioSpec beadbeater (BioSpec Products, Bartlesville, OK) for one minute. We then extracted total nucleic acids with a BioMerieux NucliSENS extraction kit with magnetic bead capture (BioMerieux, Durham, NC) following the manufacturer’s protocol.

### SARS-CoV-2 reverse transcriptase droplet digital PCR quantification and correlation with countywide COVID-19 cases

We used one-step reverse transcription droplet digital PCR (RT-ddPCR) to quantify the N1 region of the SARS-CoV-2 N gene with primer and probe sequences designed by the CDC (63, 64) and we quantified the bovine coronavirus using previously designed primers (63, 65). We set up RT-ddPCR reactions following the manufacturer’s instructions on a Bio-Rad Qx200 (Bio-Rad, Hercules, CA). For all assays, a minimum of two reactions and a total of ≥20,000 droplets were generated per sample and at least five no template control (NTC) reactions and two positive control reactions were run per 96-well plate as well as extraction-specific NTCs. Each sample was required to have a minimum of three positive droplets (66) to be included in further analyses. We also assessed RNA recovery using the BoCoV exogenous control, and samples with < 3% recovery were excluded from further analyses.

We used the SARS-CoV-2 ddPCR quantification data and compared viral loads with county-level COVID-19 reported case data from the State of California Health and Human Services Agency (https://data.chhs.ca.gov/dataset/covid-19-time-series-metrics-by-county-and-state). We calculated rolling 7-day averages for COVID-19 cases in Los Angeles, Orange, and San Diego counties with the R package “zoo”(67), and ran Pearson correlations between viral load and reported COVID-19 cases with the R package “Hmisc” (68) at each time point where we had both data points.

### Wastewater sample processing for metatranscriptomic sequencing

We followed a similar protocol as Crits-Cristoph et al 2021 and Wu et al 2020 (23, 46) to concentrate viruses and extract RNA. We pasteurized 50 mL of wastewater in a 65 °C water bath for 90 minutes, then filtered the samples through a sterile 0.22 um vacuum filter (VWR, Radnor, PA) to remove solids. We then concentrated the filtrate through ultracentrifugation at 3,000xg with 10kDa Amicon filters (MilliporeSigma, Burlington, MA) by successively centrifuging then discarding the flowthrough until the entire 50 mL sample was processed. This resulted in final volumes of < 500uL for each sample, which we then stored at -80 °C until RNA extraction. We thawed the wastewater concentrate on ice, then used an Invitrogen PureLink RNA Mini Kit with DNase (Invitrogen, Waltham, MA) to extract RNA by following the manufacturer’s protocol, quantified the resulting RNA with an AccuBlue Broad Range RNA Quantification kit (Biotium, Fremont, CA) kit on a DeNovix QFX Fluorometer (DeNovix, Wilmington, DE) spectrophotometer, then stored the RNA at -80 °C.

### Next generation sequencing library preparation

Sample library preparation and next-generation sequencing was performed by the UC Irvine Genomics High Throughput Facility (GHTF). The GHTF used two separate library preparation strategies per sample: One library was prepared with the Illumina RNA Prep with Enrichment kit (Illumina, San Diego, CA), and the other was prepared using the same preparation kit with the addition of the Illumina Respiratory Virus Oligo Panel to enrich for human respiratory viruses. The GHTF then sequenced the paired-end libraries either 2x100bp or 2x150bp (see supplemental file SF4) on an Illumina NovaSeq 6000 with an S4 300 cycle kit and sent the data as demultiplexed FASTQ files.

### Bioinformatics and sequence data processing

We used the UC Irvine High Performance Community Computing Cluster (HPC3) for all data processing on the provided FASTQ files. We removed primers, adapter sequences, and low-quality bases with “bbduk” in the BBTools software package (69), then removed PCR duplicates with BBTools “dedupe.” We downloaded the NCBI Virus RefSeq Genome database and used Bowtie2 (70) to build an index of the viral genomes, then mapped the cleaned FASTQ reads against this index. We used “Samtools” (71) to convert the resulting SAM files to sorted BAM files and finally used “inStrain” (72) to calculate all viral abundances and profile viral variants on read pairs with >90% average nucleotide identity to their respective reference genomes. Lastly, we used “mosdepth” (73) and “bedtools” (74) to calculate genome coverage and breadth across all SARS-CoV-2 sequences.

For community diversity analyses, we tabulated viral abundances, then normalized the reads into within-sample relative abundances in R (75). We used this table to generate Shannon Diversity indices and Bray-Curtis dissimilarity matrices with the R package “vegan” (76). We also ran Adonis PERMANOVA tests on the distance matrices, and performed non-metric multidimensional scaling on the data, compared the proportional abundances of viruses with ANCOM (77), and compared the relative abundances of viruses over time with “lmerTest” using WTP as a random effect (78). Lastly, we plotted all figures with “ggplot2” (79), “gggenes” (80), or “pheatmap” (81).

### Data Availability

Raw sequencing data has been deposited on the NCBI Sequence Read Archive under accession number PRJNA729801, and representative code can be found at github.com/jasonarothman/wastewater_viromics_sarscov2.

## Data Availability

Raw sequencing data has been deposited on the NCBI Sequence Read Archive under accession number PRJNA729801

https://www.ncbi.nlm.nih.gov/bioproject/PRJNA729801

## Acknowledgements

We would like to thank the staff of City of Los Angeles Sanitation and Environment, Los Angeles County Sanitation District, Orange County Sanitation District, and City of San Diego Public Utilities for collecting influent. We would also like to thank E. Macias for assistance with samples. This research was supported by Emergency COVID-19 Research Seed Funding through the University of California Office of the President Research Grants Program Office (awards #R01RG3732 and #R00RG2814) awarded to JAR, TBL, and KLW, and a Hewitt Foundation for Biomedical Research postdoctoral fellowship to JAR. This work was made possible, in part, through access to the Genomics High Throughput Facility Shared Resource of the Cancer Center Support Grant (P30CA-062203) at the University of California, Irvine and NIH shared instrumentation grants 1S10RR025496-01, 1S10OD010794-01, and 1S10OD021718-01, and access to computing resources from the UCI High Performance Cloud Computing Center.

