## Supplemental information for "RNA viromics of Southern California wastewater and detection of SARS-CoV-2 single nucleotide variants"

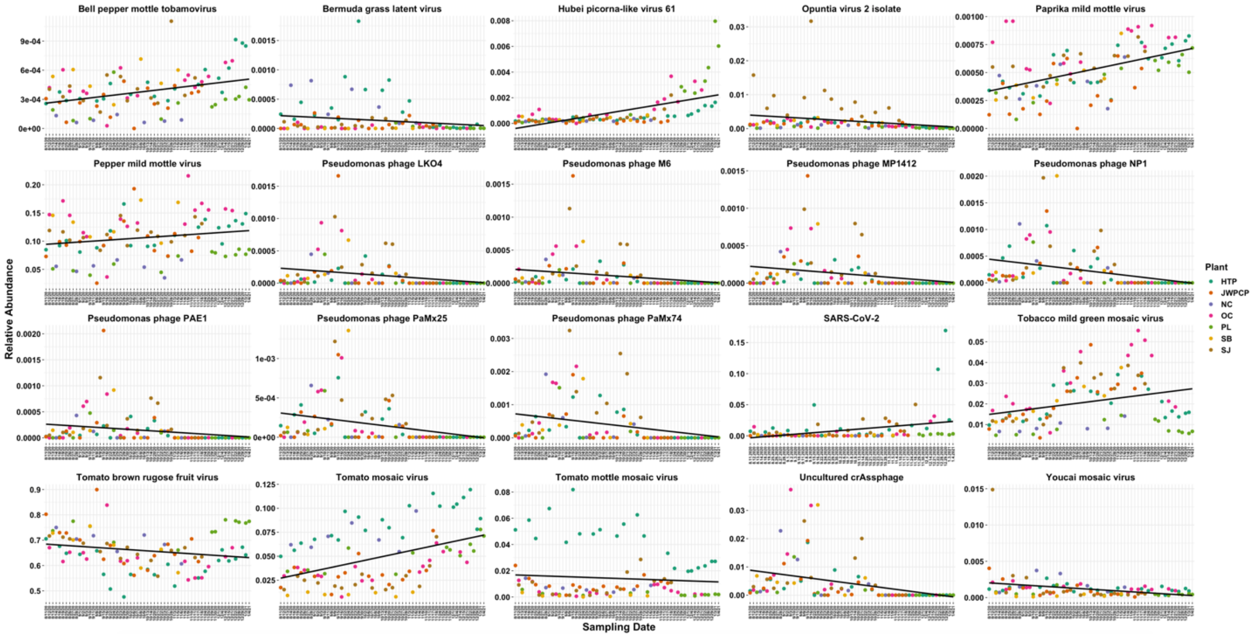


Figure S1: Relative abundances of viruses in unenriched samples (except note below) that significantly changed over time (LME $P_{adj} < 0.05$) colored by WTP. Sample dates run left to right (oldest to most recent) along the "x" axis. Note, that SARS-CoV-2 was measured and tested in enriched samples only (LME $P_{adj} < 0.05$).

Supplemental File SF1: SARS-CoV-2 RT-ddPCR quantification data, county-reported daily COVID-19 counts, and seven-day rolling averages for each sample.

Supplemental File SF2: Genomic coverage, breadth, and descriptive statistics of SARS-CoV-2 sequences for each sample.

Supplemental File SF3: Sequencing depth and Single Nucleotide Variant analyses for SARS-CoV-2 per sample after masking potentially problematic sites.

Supplemental File SF4: Sample metatranscriptomic sequencing metadata.
